# Study protocol for a factorial randomized trial of behavioral interventions to increase health care proxy documentation among older and seriously ill adults

**DOI:** 10.64898/2026.09.02.26362062

**Authors:** Jonathan N. Cloughesy, Tom Y. Chang, Susan Enguidanos, Mireille Jacobson

## Abstract

**Introduction:** Health care proxies serve as surrogate decision-makers for patients with limited decisional capacity, yet only one in three U.S. adults have designated one. This trial will evaluate low-cost, scalable behavioral interventions embedded in previsit mailed outreach to increase health care proxy documentation.

**Methods and analysis:** In this 2 × 2 + 1 factorial superiority trial, approximately 20,000 patients with an upcoming in-person primary care visit at a Northern California health system will be randomized over a 12-month period into one of four intervention mailers encouraging designation of a health care proxy, or to no mailer under usual care. Eligible patients will have no documentation of a health care proxy in the electronic health record (EHR) and will be aged 55 years or older, or 18 years or older with cancer, congestive heart failure, or chronic kidney disease. The four mailers differ by the presence or absence of a pre-commitment prompt, and whether the mailer is sent from the patient’s primary care provider or the health system. The primary outcome is EHR-documentation of a health care proxy by 30 days after the index scheduled visit date.

Secondary outcomes include proxy documentation through 90 days, legal advance care planning documentation through 30 and 90 days, and visit attendance. We will use logistic regression to estimate the intention-to-treat effect of receiving any intervention mailer versus no mailer, a pre-commitment mailer versus a mailer without pre-commitment, and a provider-signed versus health system-signed mailer.

**Ethics and dissemination:** The University of Southern California Institutional Review Board approved the protocol and granted a waiver of informed consent for this minimal-risk trial. Results will be posted on ClinicalTrials.gov within one year of the primary completion date.

**Trial Registration:** The trial was prospectively registered on December 12, 2025 at ClinicalTrials.gov (<u>NCT07290478</u>).

**Strengths and limitations of this study:**

- This individually randomized 2 × 2 + 1 factorial trial evaluates two low-cost behavioral strategies embedded within mailed outreach to increase health care proxy documentation.
- Use of routinely collected electronic health record data and a waiver of informed consent reduce consent-related selection bias, permit enrollment of the entire eligible population, and limit missing outcome data due to loss to follow-up.
- The primary outcome is health care proxy documentation in the electronic health record; however, the trial does not assess patient-proxy communication, proxy preparedness, goal-concordant care, or other downstream clinical outcomes.
- The trial is conducted within a large, primarily Medicaid-serving integrated health system in Northern California. Results of this trial may not be generalizable to other populations, health care systems, and jurisdictions with different health care proxy designation requirements.

## Introduction

Designation of a health care proxy—a person authorized to make medical decisions on behalf of a patient with limited decisional capacity—is a foundational component of patient-centered care and advance care planning.^1^ Having a designated proxy is associated with receiving care that is aligned with patients’ goals and reduced proxy distress, anxiety, and grief.^2,3^ Loss of decisional capacity is common: estimates suggest one-quarter of hospitalized adults lack medical decision-making capacity at any given time,^4^ and many older adults and those with serious illness will require proxy decision-making at the end of life.^5^ Despite this, only about one in three U.S. adults have a designated health care proxy.^6^ The gap between need and documentation represents a missed opportunity to support goal-concordant care and improve outcomes for seriously ill adults and their caregivers.

While much of the literature has focused broadly on advance care planning (ACP),^3,7,8^ some critics contend that ACP has not consistently improved end-of-life outcomes, in part because people cannot reliably anticipate their future preferences in the context of serious illness.^9–13^ Even critics, however, point to the importance of establishing a trusted decision-maker who can interpret and communicate a patient’s values in real time.^12,14^ Compared with prespecified treatment directives, proxy designation offers flexibility in the face of clinical uncertainty. Yet, despite broad conceptual support, health care proxy documentation remains underutilized.

The reasons for low levels of health care proxy documentation are multifaceted and not well understood. While some research points to knowledge deficits,^15,16^ initiatives to improve patient knowledge have not meaningfully increased proxy designation.^17^ In most states, legal designation requires the signature of two qualified witnesses or acknowledgment by a notary public,^18^ creating significant logistical, administrative, and psychological burdens that may discourage completion.^19^ Time constraints during clinical encounters and low clinician efficacy further limit clinician-initiated discussions,^20^ and patients similarly report lack of time, reluctance to initiate, and procrastination as barriers to taking action.^21,22^ However, most patients are able and willing to identify a health care proxy when asked to do so.^23^ And, in many states including California, oral health care proxy designation is permitted under limited circumstances, such as when made during the course of treatment at a health care facility.^24^ This suggests that low levels of health care proxy documentation may not reflect resistance or uncertainty, but rather behavioral frictions that impede follow-through.

Behavioral economic theory points to present bias, the tendency to overweight immediate costs relative to delayed benefits,^25–27^ as a possible reason for low proxy documentation. Designating a health care proxy entails immediate effort (reviewing forms, discussions with loved ones, returning documentation), while its benefits are uncertain and temporally distant. As a result, even individuals who endorse the value of proxy documentation may delay action indefinitely. Interventions that acknowledge these behavioral barriers may therefore meaningfully increase uptake.

Pre-commitment prompts, a behavioral strategy that asks individuals to commit in the present to complete a beneficial but effortful action in the future, may be a useful tool to increase health care proxy documentation. Pre-commitment prompts substitute a high-effort behavior (e.g., proxy designation) for a low-effort action (e.g., a commitment to designate), and leverage individuals’ desire for consistency with prior commitments to mitigate present bias and increase completion of the desired behavior.^28,29^ While widely studied in other behavioral domains, this approach has not been tested to promote health care proxy documentation.

Message source may also influence health care proxy documentation. Individuals are more responsive to recommendations delivered by trusted sources,^30^ and the vast majority of Americans report high levels of trust in their own health care providers.^31^ Prior studies show that referral recommendations, such as those for palliative care, are more acceptable when originating from a patient’s primary care provider.^32^ While patients and providers may both be unlikely to initiate discussions about health care proxy documentation during visits, pre-visit written requests that are signed by a patient’s primary care provider may be an effective way to harness the influence of a trusted messenger and create an expectation of proxy documentation at an upcoming visit.

To date, no randomized trial has evaluated behavioral strategies specifically designed to increase health care proxy documentation independent of broader advance care planning interventions. There remains a need for scalable, low-cost interventions that can be deployed upstream of clinical encounters to increase proxy documentation. This trial addresses that gap by testing pre-commitment prompts and provider requests embedded within mailed communications sent prior to scheduled primary care visits.

### Objectives

The primary objective of this superiority trial is to evaluate whether behavioral strategies embedded within mailed communication sent prior to a scheduled primary care visit increase health care proxy documentation in the electronic health record by 30 days after the scheduled visit date. The trial will include participants without a documented health care proxy who are aged 55 years or older, or 18 years or older and have cancer, congestive heart failure, or chronic kidney disease.

Eligible participants with an upcoming primary care visit will be randomized to receive one of four mailers that vary according to two behavioral interventions—the presence or absence of a pre-commitment prompt and whether the mailer is signed by the patient’s primary care provider or the health system—or to receive no mailer under usual care.

The trial has three primary estimands. First, among all randomized participants, we will estimate the effect of assignment to an intervention mailer compared with no mailer. Second, among participants randomized to the four mailer arms, we will estimate the average effect of assignment to a pre-commitment prompt versus no pre-commitment prompt, averaging equally across provider-signed and health system-signed mailers. Third, among participants randomized to the mailer arms, we will estimate the average effect of assignment to a provider-signed versus health system-signed mailer, averaging equally across mailers with and without a pre-commitment prompt.

The trial’s secondary objectives are to evaluate intervention effects on health care proxy documentation by 90 days and on documentation of any legal advance care planning document by 30 and 90 days after the scheduled primary care visit. To assess potential unintended effects, we will also evaluate whether assignment to an intervention mailer affects attendance at the originally scheduled visit or attendance at any primary care visit within 90 days.

The trial is prospectively registered on ClinicalTrials.gov (<u>NCT07290478</u>). This protocol adheres to the Standard Protocol Items: Recommendations for Interventional Trials (SPIRIT) 2025 reporting guidelines^33,34^ and the SPIRIT extension for factorial trials.^35,36^

## Methods and analysis

### Trial design and setting

This trial employs a 2 × 2 + 1 factorial randomized superiority design to evaluate the impact of two behavioral interventions on health care proxy documentation. Participants will be randomized at the individual-level in a target 1:1:1:1:1 allocation ratio to one of the four factorial mailer arms or the no-mailer control arm. Randomization will be stratified by age group (18–54, 55–64, or ≥65 years) and inclusion on a qualifying disease registry (yes or no). The factorial design was chosen to efficiently estimate the average effects of each behavioral intervention.

The trial will take place at Contra Costa Health (CCH), a primarily Medicaid-serving health system with nine outpatient primary care clinics located throughout Contra Costa County in Northern California. Patients will be considered eligible for the trial if they do not have a health care proxy documented in the EHR, and are either aged 55 years and older, or aged 18 years and older and on a CCH patient registry for cancer, chronic kidney disease, or congestive heart failure. Patients without an assigned primary care provider for their scheduled appointment will be excluded before randomization. Eligible patients with a scheduled in-person visit with a primary care provider at an outpatient primary care clinic will be identified from EHR data and enrolled in the trial under a waiver of informed consent. The waiver improves the generalizability of the results by avoiding consent-related selection into the trial and permitting enrollment of the full eligible population. Patients will be assessed for eligibility for visits scheduled during a 12-month period. Each patient will be randomized only once. For patients with multiple qualifying scheduled visits, the first scheduled visit that results in randomization will be designated the index scheduled visit date, and the patient will not be eligible for subsequent enrollment.

### Patient and public involvement

No patient or public involvement is planned in the design, conduct, or reporting of the trial.

### Intervention and comparator arms

Patients allocated into the intervention arms will be sent a mailer to their home address, while participants allocated to the control arm will not be sent a mailer, and will continue to receive usual care without modification. The intervention mailer contains a letter describing the importance of designating a health care proxy, a list of frequently asked questions, a form to designate up to two health care proxies, and a request to bring the completed health care proxy form to the patient’s next visit. The completed form is intended to facilitate, rather than itself constitute, designation of a health care proxy; patients are asked to bring it to their next visit to facilitate oral designation and documentation in the EHR. The mailer does not serve as a legally effective designation. The mailer is written in English, and a Spanish language version will be sent to participants who indicated a Spanish language preference at CCH. The intervention mailers are presented in **Supplementary Materials**.

The mailer differs across the 2 × 2 factorial component, which crosses: (a) the source of the messenger (patient’s primary care provider or the health system), and (b) the presence or absence of a pre-commitment prompt to bring the completed form to the patient’s next visit. Intervention materials will be printed and mailed by health system staff. Monitoring of the print and mail process will be conducted intermittently by trial personnel to ensure adherence to protocol. Because the intervention consists of low-risk mailed communication, there are no pre-specified criteria for modification or discontinuation of the intervention.

A pre-existing health system intervention will occur during the trial for patients aged 65 and above without a documented health care proxy. While in the waiting room prior to their visit, these patients may receive a brief letter explaining the importance of a health care proxy and be provided with a form to designate one or more health care proxies. Patients aged 65 and older across all trial arms will be exposed to the co-occurring intervention.

### Participant timeline

Every other week, the study team will identify patients who meet all eligibility criteria and have an in-person primary care visit scheduled 7–21 days later. Eligible patients will then be randomly assigned to a study arm. For patients assigned to an intervention arm, a corresponding mailer will be printed and mailed approximately 7–21 days prior to the scheduled visit. Outcome data will be obtained from routinely collected EHR documentation. All data is collected as part of routine clinical care, and no additional study visits or research-specific data collection will occur. A schematic diagram of the participant timeline is presented in **Figure 1**.

**Figure 1.**
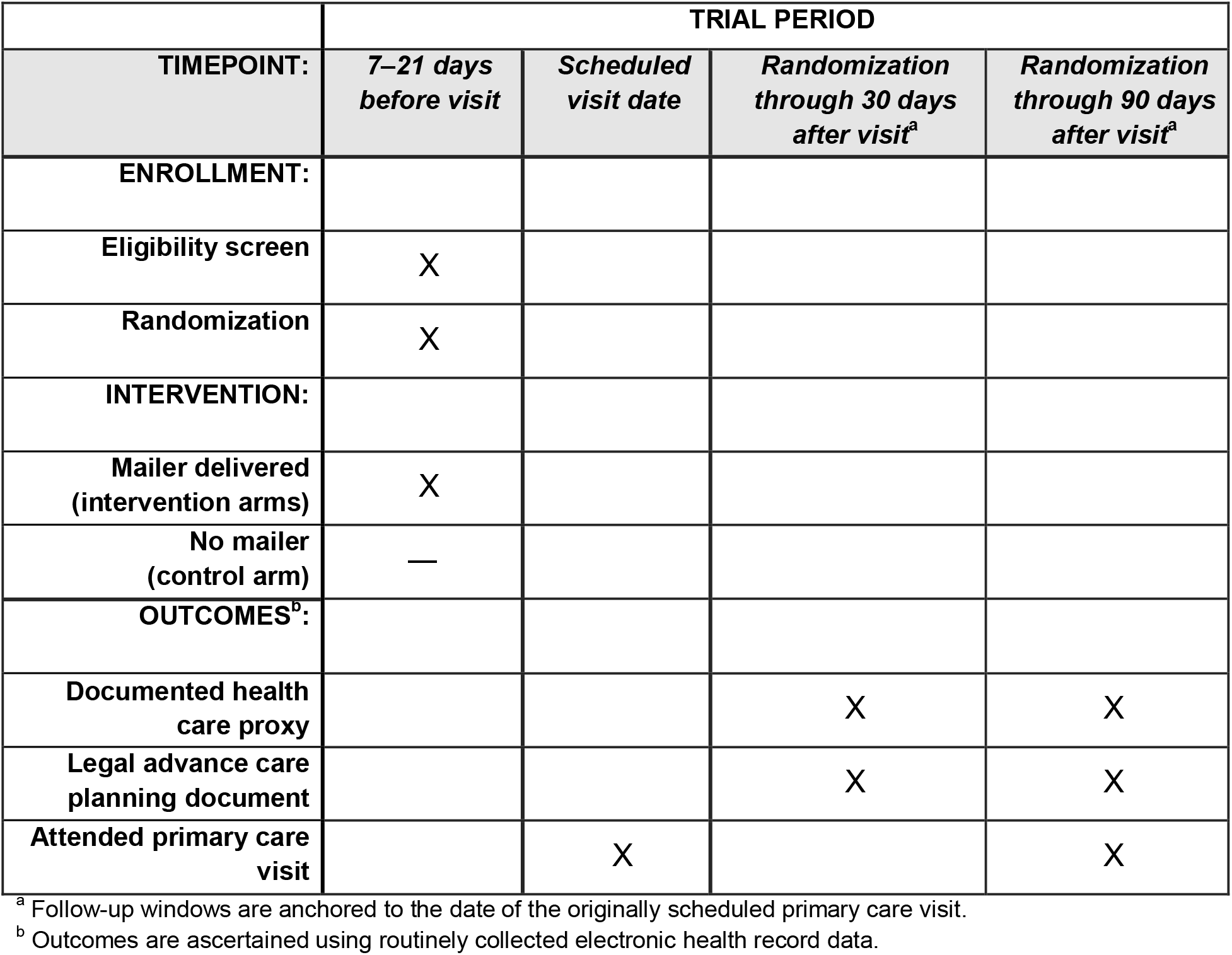
Participant timeline.

### Outcomes

The primary outcome is the proportion of patients with a health care proxy documented in the EHR at any time from randomization through 30 days after the patient’s index scheduled primary care visit date. The outcome timepoint will not be adjusted if the originally scheduled visit date is cancelled or rescheduled. Documentation of a health care proxy in the EHR was chosen as the primary outcome because the EHR is frequently consulted when determining who the patient prefers as a health care proxy. Making proxy information easily accessible to providers in the EHR can facilitate care that aligns with patient preferences across health care settings.

Secondary outcomes include the proportion of patients with health care proxy documentation in the EHR from randomization through 90 days after the index scheduled visit date, and the proportion of patients with any legal advance care planning documentation, including advance directives or power of attorney, in the EHR from randomization through 30 and 90 days after the index scheduled visit date. Given the minimal-risk nature of the intervention, no systematic collection of adverse events is planned. While no adverse events are expected to occur, the intervention may result in some patients experiencing discomfort, leading to visit cancellation.

To evaluate this possibility, the proportion of patients attending their index scheduled visit, as well as the proportion of patients attending a primary care visit from randomization through 90 days after the index scheduled visit date will be assessed. The research team will correspond with health system leadership throughout the study period to assess any unexpected safety concerns.

## Statistical analysis

Our statistical analysis will use an adjusted logistic regression model containing four indicator variables for the four factorial mailer arms, with the no-mailer arm as the reference group. Analyses will follow the intention-to-treat principle by including all participants according to their randomized assignment regardless of mailer receipt, scheduled visit attendance, or other intercurrent events.^36^

The model will adjust for the randomization strata defined by age group and qualifying disease-registry status, the randomization batch, the days between randomization and the scheduled visit, and fixed effects for the assigned primary care provider at randomization. Additional baseline covariates will include continuous age, sex, race and Hispanic ethnicity, and primary insurer. Covariates are intended to improve precision and account for the randomization procedure. Patients with missing baseline covariate data will remain in analyses. For continuous covariates, missing values will be imputed using the pooled mean across all randomized participants, and a corresponding missingness indicator will be included in the model. For categorical covariates, missing or unknown values will be included as a separate category. Adjusted and unadjusted analyses will be reported.

Outcome probabilities will be estimated for each arm using marginal standardization over the empirical distribution of baseline covariates. Absolute treatment effects will be summarized using risk differences with 95% confidence intervals. Inference will rely on two-sided tests of the risk-difference contrasts, with the significance level set to α = .05. We will also report relative effect sizes using risk ratios with 95% confidence intervals. Because treatment is randomized at the individual level, confidence intervals and hypothesis tests will use heteroskedasticity-robust standard errors.^37^ We will also calculate standard errors clustered at the provider level as a sensitivity analysis.

The effect of the intervention mailer will be estimated as the equal-weight average of the probabilities in the four mailer arms minus the probability in the no-mailer arm. The pre-commitment effect will be estimated as the equally weighted average of the pre-commitment versus no pre-commitment risk differences within the provider-signed and health system-signed mailer arms. Similarly, the message-source effect will be estimated as the equally weighted average of the provider-signed versus health system-signed risk differences within the pre-commitment and no pre-commitment arms. Each of the three primary comparisons addresses a distinct research question and will be tested separately without adjustment for multiplicity; consequently, the familywise type I error rate across the three primary comparisons will not be controlled.^38^

To assess whether the average effects of each behavioral intervention conceal effect heterogeneity, we will report the pre-commitment by message-source interaction estimate on the risk-difference scale and its 95% confidence interval, the outcome probability in each of the four mailer arms, and the effect of each factor separately at each level of the other factor.^36,39–41^ For example, the effect of provider-signed versus health system-signed mailer will be reported separately for patients assigned to a mailer with a pre-commitment prompt and patients assigned to a mailer without a pre-commitment prompt.

Secondary analyses will assess whether treatment effects differ by age group (<65 versus ≥65 years) and qualifying disease registry status. To do so, we will interact the subgroup indicator with each of the four mailer-arm indicators. The complete statistical analysis plan is provided in the **Supplementary Materials**.

### Sample size

The trial will use a fixed 12-month recruitment period and will include all eligible patients identified during that period. We anticipate enrolling approximately 20,000 participants, corresponding to approximately 4,000 participants per trial arm. Because the sample size is determined by a fixed recruitment period, we conducted a simulation-based power calculation to evaluate whether the anticipated enrollment would provide adequate power for the three primary comparisons.

Statistical power was evaluated using Monte Carlo simulation with 5,000 replications. Participants were allocated equally among the five trial arms. We assumed an outcome probability of 1.0% in the no-mailer arm, and probabilities of 10.0% in the no-prompt, health system-signed arm, 11.5% in each of the two single-factor arms, and 13.0% in the prompt, provider-signed arm. These probabilities correspond to an average probability of approximately 11.5% across the four mailer arms, a 10.5-percentage-point risk difference for any mailer versus no mailer, a marginal risk difference of 1.5 percentage points for the effects of the pre-commitment prompt and provider-signed mailers, and no interaction between factors on the risk-difference scale.

Following our planned analytic strategy, we fit a logistic regression model containing four indicators for the factorial mailer arms, with the no-mailer arm as the reference group. Power was calculated as the proportion of 5,000 replications in which the null hypothesis for the risk-difference contrast was rejected, using a two-sided Wald test with heteroskedasticity-robust standard errors and a significance level of α = .05. No covariates were included in the model.

The anticipated sample size of 20,000 provided over 99% power to detect a 10.5-percentage-point difference between the combined mailer arms and no-mailer arm, and approximately 85% power to detect a 1.5-percentage-point marginal effect for each factorial comparison. A lower total sample size of 15,000 was estimated to provide over 99% power for the mailer versus no-mailer comparison, and over 72% power for each factorial comparison.

### Assignment to arms

#### Sequence generation

A member of the study team (T.Y.C.) will generate random treatment assignments using the Stata randtreat command.^42^ Participants will be assigned in approximately equal proportions to the five study arms within strata, including age group (18–54, 55–64, or ≥65 years) and qualifying disease-registry status (yes or no). When the number of participants in a stratum is not divisible by five, the remaining participants will be randomly allocated among the five arms within that stratum, such that the numbers assigned to the arms within each stratum differ by no more than one.

#### Allocation concealment mechanism

Patients included in each biweekly randomization batch will be finalized before the randomization program is executed. All participants in the batch will then be randomized simultaneously using the method described above. Treatment assignments will not be available to study personnel before randomization; therefore, knowledge of allocation cannot influence eligibility determination or participant inclusion.

#### Implementation

The study team member (T.Y.C.) implementing the computer-generated randomization will allocate each participant into study arms based on the results of the randomization. The study team will then send the list of patients allocated to intervention arms to CCH staff, who will print and mail the appropriate mailer for each enrolled patient based on their arm assignment. Participants assigned to the no-mailer control arm will receive usual care and will not appear on the mailing list.

#### Blinding

This trial is considered open-label as patients cannot be blinded to allocation because they will be delivered the intervention based on their arm assignment. Although providers will not be directly informed of the patient’s arm assignment, the patient is encouraged to bring their mailer to the appointment which may inform the provider of the patient’s assignment. Data analysts will not be blinded to arm assignment during analysis.

### Data collection, management, and analysis

#### Data collection methods

Data will be collected electronically in the CCH EHR. A de-identified report will be generated containing relevant data fields and transmitted to the study team.

#### Data management

Electronic data capture minimizes errors and missing data, improving the robustness of data collection. Data will be stored at CCH until the end of the trial, when it will be transferred to the study team.

### Monitoring

#### Data monitoring committee

A data and safety monitoring board (DSMB) will conduct monitoring of trial activities. The DSMB consists of an independent group of experts that provides recommendations to the study investigators and the funding agency. The DSMB comprises members from various backgrounds including Clinical & Behavioral Health, Public Health & Epidemiology, Biostatistics & Data Science, Clinical Medicine & Nursing, Neuroscience & Sleep Research, and Healthcare Systems, Policy & Ethics. There are no interim analyses or stopping guidelines planned for this trial due to the absence of any expected adverse events. The DSMB charter is presented in the **Supplementary Materials**.

#### Trial monitoring

Systematic trial monitoring is not planned. The study team will be in frequent communication with health system staff in charge of distributing intervention materials, as well as advance care planning leadership.

### Ethics and dissemination

#### Research ethics approval

The trial protocol was approved by the University of Southern California Institutional Review Board (UP-23-01203) on January 19, 2024.

#### Protocol amendments

Substantive amendments to the trial protocol will be communicated to the University of Southern California Institutional Review Board, the DSMB, and the funding agency.

#### Consent or assent

A waiver of informed consent was obtained for participant enrollment because the research was determined to involve no more than minimal risk to participants, and because a waiver would not adversely affect the rights and welfare of subjects.

#### Confidentiality

All identifiers, including patient name, date of birth, and medical record number will be removed before being shared with the study team.

#### Dissemination policy

Trial results will be posted on the study’s ClinicalTrials.gov record within one year of the primary completion date, and will be submitted for publication in a peer-reviewed journal.

#### Data sharing

De-identified individual participant-level data and the analytic code used for results publication may be shared upon request and with approval by the health system, as governed by a data use agreement signed by both parties.

#### Ancillary and post-trial care

There are no plans for ancillary or post-trial care, as it is not deemed necessary in this trial.

## Discussion

The proposed trial will evaluate low-cost, behaviorally informed interventions designed to increase health care proxy documentation among older adults and adults with serious illness. Specifically, the trial will test whether mailed communication sent in advance of a scheduled primary care visit can prompt patients to designate a health care proxy. Using a 2 × 2 + 1 design, the intervention will vary whether the mailer includes a pre-commitment prompt encouraging patients to designate a proxy at their next visit, and the signatory and thus proposed messenger for the mailer—either the health system or the patient’s own primary care provider. An additional control group will receive usual care with no mailed outreach.

This design will allow us to estimate the effects of behavioral pre-commitment and message source, while also benchmarking the combined effect of the mailers against usual care. By intervening before a routine clinical encounter, the mailer is intended to increase salience, generate commitments, and build expectations of follow-through at a moment when patients are already engaged with the health care system.

If effective, this intervention would offer a scalable and easily implementable strategy to increase health care proxy documentation. The approach requires minimal staff time, can be automated within existing appointment workflows, and relies on materials that can be readily adapted across settings. As such, it has the potential to be deployed broadly within health systems with low or incomplete documentation rates, including safety-net systems serving vulnerable populations, thereby improving patient preparedness and supporting patient-centered care.

## Supporting information

Supplementary Materials: Intervention Mailers

Supplementary Materials: Statistical Analysis Plan

Supplementary Materials: DSMB Charter

Supplementary Materials: SPIRIT Checklist

## Data Availability

No data is used in this study.

## Limitations

This trial has a number of limitations. First, we do not measure process outcomes including mailer receipt, patient-proxy communication, and proxy preparedness, nor goal-concordant care or other downstream clinical outcomes. Second, patient-level randomization may result in spillovers between conditions if provider exposure to an intervention arm changes behavior toward patients assigned to other trial arms. Third, the trial may have limited generalizability to other populations, health systems, or jurisdictions with different requirements for health care proxy designation. Because the intervention mailer does not serve as a legal document, the interventions may not be applicable in jurisdictions that do not accept oral proxy designations. Fourth, the trial is not designed to detect an interaction between interventions. However, it is possible that each intervention will have different effects depending on the level of the other factor.

### Abbreviations

ACP: Advance care planning
CCH: Contra Costa Health
DSMB: Data and Safety Monitoring Board
EHR: Electronic health record
SPIRIT: Standard Protocol Items: Recommendations for Interventional Trials

## Declarations

### Author contributions

**Jonathan N. Cloughesy:** Conceptualization, Methodology, Validation, Formal Analysis, Investigation, Resources, Writing - Original Draft, Writing - Review & Editing; **Tom Y. Chang:** Conceptualization, Methodology, Validation, Investigation, Resources, Data Curation, Writing - Review & Editing, Project Administration; **Susan Enguidanos:** Conceptualization, Methodology, Validation, Investigation, Resources, Writing - Original Draft, Writing - Review & Editing; **Mireille Jacobson:** Conceptualization, Methodology, Validation, Investigation, Resources, Writing - Original Draft, Writing - Review & Editing, Supervision, Project Administration, Funding Acquisition

### Funding

This work was supported by the National Institute on Aging grant number P30AG024968. The National Institute on Aging had no role in the design, implementation, or analysis of this study.

### Competing interests statement

The authors declare no competing interests.

## Acknowledgments

We thank Julie Freedman, Duane Eikleberry, Contra Costa Health Print & Mail, Tara Knight, & Lisa Mendelow for their invaluable assistance with the conduct of this trial.

## Notes

### Competing Interest Statement

The authors have declared no competing interest.

### Clinical Trial

NCT07290478

### Author Declarations

IRB of University of Southern California gave ethical approval for this work on January 19, 2024 (UP-23-01203).

