## Supplementary Materials: Intervention Mailers for "Study protocol for a factorial randomized trial of behavioral interventions to increase health care proxy documentation among older and seriously ill adults"

Provider-signed mailer with pre-commitment prompt

CONTRA COSTA HEALTH

### Choosing your medical decision-maker

**Dear <<First Name>>:**

I'm looking forward to your upcoming visit on <<date>>. Before you come in, I would like you to consider an important question.

If you ever become too sick to make your own medical decisions, is there someone you trust who could make them for you? Sharing this information will help us take the best possible care of you.

A good medical decision-maker is someone who knows you well and understands what is most important to you. In an emergency, they would ask doctors questions and make choices that respect your wishes.

You can choose one or more people. Please write down their name(s) on the other side of this page and bring this form to your next appointment.

Will you write your decision-maker's name on this form and hand it to me at your next visit?

☐ I, <<First Last>>, commit to naming my decision-maker at my next visit on <<date>>.

If you have any questions, we can discuss them at your visit.

Sincerely,

**<<First Name>> <<Last Name>>, <<Degree>>**

FREQUENTLY ASKED QUESTIONS:

#### What is a medical decision-maker?

If you're in an emergency or become very sick and can't speak for yourself, a **medical decision-maker** is someone you trust to make medical decisions for you.

#### What can they decide?

- Where you get care
- Which treatments you receive
- Whether you get life support (CPR, breathing machine, dialysis, feeding tube)
- What happens to your body after death

#### Why is this important?

About 3 in 4 people will face a time when they can't make their own medical decisions. If you don't choose someone, state law or doctors may choose for you.

#### Who should you choose?

**Someone 18 or older who:**

- Can talk with you about your health and medical care
- Understands your wishes and priorities
- Will speak up for you and manage disagreements

Make sure to ask if they're willing to take on this role.

#### Other tips:

- Talk regularly with your decision-maker about your wishes.
- Name a backup in case your first choice is unavailable.
- You can change your decision-maker at any time.

Turn over to choose your medical decision-maker →

Health system-signed mailer with pre-commitment prompt

CONTRA COSTA HEALTH

### Choosing your medical decision-maker

**Dear <<First Name>>:**

We're looking forward to your upcoming visit on <<date>>. Before you come in, we would like you to consider an important question.

If you ever become too sick to make your own medical decisions, is there someone you trust who could make them for you? Sharing this information will help us take the best possible care of you.

A good medical decision-maker is someone who knows you well and understands what matters most to you. In an emergency, they would ask doctors questions and make choices that respect your wishes.

You can choose one or more people. Please write down their name(s) on the other side of this page and bring this form to your next appointment.

Will you write your decision-maker's name on this form and hand it to your provider at your next visit?

☐ I, <<First Last>>, commit to naming my decision-maker at my next visit on <<date>>.

If you have any questions, you can discuss them at your visit.

Sincerely,

**Contra Costa Health**

FREQUENTLY ASKED QUESTIONS:

#### What is a medical decision-maker?

If you're in an emergency or become very sick and can't speak for yourself, a **medical decision-maker** is someone you trust to make medical decisions for you.

#### What can they decide?

- Where you get care
- Which treatments you receive
- Whether you get life support (CPR, breathing machine, dialysis, feeding tube)
- What happens to your body after death

#### Why is this important?

About 3 in 4 people will face a time when they can't make their own medical decisions. If you don't choose someone, state law or doctors may choose for you.

#### Who should you choose?

**Someone 18 or older who:**

- Can talk with you about your health and medical care
- Understands your wishes and priorities
- Will speak up for you and manage disagreements

Make sure to ask if they're willing to take on this role.

#### Other tips:

- Talk regularly with your decision-maker about your wishes.
- Name a backup in case your first choice is unavailable.
- You can change your decision-maker at any time.

Turn over to choose your medical decision-maker →

Provider-signed mailer without pre-commitment prompt

CONTRA COSTA HEALTH

### Choosing your medical decision-maker

**Dear <<First Name>>:**

I'm looking forward to your upcoming visit on <<date>>. Before you come in, I would like you to consider an important question.

If you ever become too sick to make your own medical decisions, is there someone you trust who could make them for you? Sharing this information will help us take the best possible care of you.

A good medical decision-maker is someone who knows you well and understands what is most important to you. In an emergency, they would ask doctors questions and make choices that respect your wishes.

You can choose one or more people. Please write down their name(s) on the other side of this page and bring this form to your next appointment.

If you have any questions, we can discuss them at your visit.

Sincerely,

**<<First Name>> <<Last Name>>, <<Degree>>**

FREQUENTLY ASKED QUESTIONS:

#### What is a medical decision-maker?

If you're in an emergency or become very sick and can't speak for yourself, a **medical decision-maker** is someone you trust to make medical decisions for you.

#### What can they decide?

- Where you get care
- Which treatments you receive
- Whether you get life support (CPR, breathing machine, dialysis, feeding tube)
- What happens to your body after death

#### Why is this important?

About 3 in 4 people will face a time when they can't make their own medical decisions. If you don't choose someone, state law or doctors may choose for you.

#### Who should you choose?

**Someone 18 or older who:**

- Can talk with you about your health and medical care
- Understands your wishes and priorities
- Will speak up for you and manage disagreements

Make sure to ask if they're willing to take on this role.

#### Other tips:

- Talk regularly with your decision-maker about your wishes.
- Name a backup in case your first choice is unavailable.
- You can change your decision-maker at any time.

Turn over to choose your medical decision-maker →

Health system-signed mailer without pre-commitment prompt

CONTRA COSTA HEALTH

### Choosing your medical decision-maker

**Dear <<First Name>>:**

We're looking forward to your upcoming visit on <<date>>. Before you come in, we would like you to consider an important question.

If you ever become too sick to make your own medical decisions, is there someone you trust who could make them for you? Sharing this information will help us take the best possible care of you.

A good medical decision-maker is someone who knows you well and understands what matters most to you. In an emergency, they would ask doctors questions and make choices that respect your wishes.

You can choose one or more people. Please write down their name(s) on the other side of this page and bring this form to your next appointment.

If you have any questions, you can discuss them at your visit.

Sincerely,

**Contra Costa Health**

FREQUENTLY ASKED QUESTIONS:

#### What is a medical decision-maker?

If you're in an emergency or become very sick and can't speak for yourself, a **medical decision-maker** is someone you trust to make medical decisions for you.

#### What can they decide?

- Where you get care
- Which treatments you receive
- Whether you get life support (CPR, breathing machine, dialysis, feeding tube)
- What happens to your body after death

#### Why is this important?

About 3 in 4 people will face a time when they can't make their own medical decisions. If you don't choose someone, state law or doctors may choose for you.

#### Who should you choose?

**Someone 18 or older who:**

- Can talk with you about your health and medical care
- Understands your wishes and priorities
- Will speak up for you and manage disagreements

Make sure to ask if they're willing to take on this role.

#### Other tips:

- Talk regularly with your decision-maker about your wishes.
- Name a backup in case your first choice is unavailable.
- You can change your decision-maker at any time.

**Turn over to choose your medical decision-maker** →

Provider-signed mailer

**Choose your medical decision-maker(s) below:**

Full name: ______________________

Relationship to you: ______________________

Phone number: ______________________

Full name: ______________________

Relationship to you: ______________________

Phone number: ______________________

### Hi <<First Name>>,
### it’s <<Provider>>,
### from Contra Costa Health.

**I have a simple request.**

CONTRA COSTA HEALTH

**<<Provider Name>>**
**Contra Costa Health**
<<Address line 1>>
<<Address line 2>>
<<City>>, <<State>> <<Zip>>

Health system-signed mailer

Choose your medical decision-maker(s) below:

Full name: ____________________

Relationship to you: ____________________

Phone number: ____________________

Full name: ____________________

Relationship to you: ____________________

Phone number: ____________________

#### Hi <<First Name>>,
#### it’s Contra Costa Health.

**We have a simple request.**

CONTRA COSTA HEALTH

**Contra Costa Health**
<<Address line 1>>
<<Address line 2>>
<<City>>, <<State>> <<Zip>>
