## Supplementary Materials: Statistical Analysis Plan for "Study protocol for a factorial randomized trial of behavioral interventions to increase health care proxy documentation among older and seriously ill adults"

| | |
|---|---|
| **Sponsor** | National Institute on Aging |
| **Grant Number** | P30AG024968 |
| **Principal Investigator** | Mireille Jacobson |
| **ClinicalTrials.gov record** | NCT07290478 |
| **USC IRB number** | UP-23-01203 |
| **Approval date** | August 27, 2026 |

**Objectives**
This superiority trial will evaluate behavioral strategies to increase health care proxy documentation at Contra Costa Health, a primarily Medicaid-serving health system in Northern California. Behavioral strategies will be embedded within mailed communications sent to patients prior to an upcoming scheduled primary care visit. Mailers will vary by the presence or absence of a pre-commitment prompt to designate a health care proxy and bring the mailer form to the patient's upcoming visit, and whether the mailer is sent by the patient's primary care provider or the health system. Eligible participants are adults without a health care proxy documented in the electronic health record (EHR) who are aged 55 years or older, or aged 18 years or older and included on a qualifying registry for cancer, congestive heart failure, or chronic kidney disease.

**Design**
Participants will be individually randomized in a 2 × 2 + 1 factorial design to receive one of four mailers or no mailer. Participants will be randomized in a 1:1:1:1:1 target allocation ratio. Randomization will occur in biweekly batches among patients with a scheduled primary care visit during the subsequent 7–21 days. Participants without an assigned primary care provider will be excluded prior to randomization. Randomization will be stratified by age group (18–54, 55–64, and ≥65 years) and inclusion on a qualifying disease registry. Qualifying disease registries include the Contra Costa Health patient registry for cancer, congestive heart failure, and chronic kidney disease. Each participant will be randomized only once.

**Estimands**
The trial has three primary estimands. First, among all randomized participants, we will estimate the effect of assignment to any intervention mailer compared with no mailer. Second, among participants randomized to the four mailer arms, we will estimate the average effect of assignment to a pre-commitment prompt versus no prompt, averaging equally across provider-signed and health system-signed mailers. Third, among participants randomized to the four mailer arms, we will estimate the average effect of assignment to a provider-signed versus health system-signed mailer, averaging equally across mailers with and without a pre-commitment prompt.

The analyses will follow the intention-to-treat principle. All randomized participants will be analyzed according to their assigned arm regardless of whether the assigned mailer was delivered or received, whether the scheduled visit was attended, and other intercurrent events.

**Outcomes**
The primary outcome is an indicator for health care proxy documentation in the EHR at any time from randomization through 30 days after the index scheduled primary care visit date. The outcome window will remain anchored to the originally scheduled visit date even if that visit is rescheduled or cancelled. An EHR record of health care proxy documentation will be coded as documentation present, and the absence of an EHR record will be coded as no documentation.

Secondary outcomes are: (1) health care proxy documentation in the EHR from randomization through 90 days after the index scheduled visit date; (2) any legal advance care planning documentation in the EHR, including an advance directive or power of attorney, from randomization through 30 days after the index scheduled visit date; (3) any legal advance care planning documentation in the EHR, including an advance directive or power of attorney, from randomization through 90 days after the index scheduled visit date; (4) attendance at the scheduled primary care visit, and (5) attendance at any primary care visit (including the originally scheduled primary care visit) from randomization through 90 days after the index scheduled visit date.

**Analytic Model**

For each binary outcome, the primary adjusted analysis will use a logistic regression model containing four indicator variables for the four factorial mailer arms, with the no-mailer arm as the reference group. Defining the pre-commitment prompt arm indicators as $P[x]$, the no prompt arm indicators as $N[x]$, the provider-signed indicators as $[x]P$, and the system-signed indicators as $[x]S$, where $Y_i$ refers to the outcome for individual $i$, the model will be fitted as:

$$Y_i = \beta_0 + \beta_1 PP_i + \beta_2 PS_i + \beta_3 NP_i + \beta_4 NS_i + \epsilon_i$$

The model will adjust for the randomization strata defined by age group and qualifying registry status, randomization batch, the number of days between randomization and the index scheduled visit, and fixed effects for the primary care provider assigned at randomization. Additional baseline patient covariates include age in years, sex, race and Hispanic ethnicity, and primary insurer. Patients with missing baseline covariate data will remain in analyses. For continuous covariates, missing values will be imputed using the pooled mean across all randomized participants, and a corresponding missingness indicator will be included in the model. For categorical covariates, missing or unknown values will be included as a separate category. We will also report an unadjusted model. Residuals captured by the error term $\epsilon$ are assumed to follow a standard logistic distribution.

**Comparisons and Inference**

For each arm, we will report the adjusted outcome probability. For each primary comparison, we will report the risk difference, risk ratio, and 95% confidence interval. Statistical significance will be assessed using two-sided tests at $\alpha = .05$. Heteroskedasticity-robust standard errors will be used for the primary analysis. As a sensitivity analysis, standard errors will also be clustered by primary care provider.

The outcome probability for each trial arm will be estimated by marginal standardization over the empirical distribution of baseline covariates in the analysis population. Defining the probabilities for no mailer arm as $pC$, the pre-commitment prompt arms as $pP[x]$, the no prompt arms as $pN[x]$, the provider-signed arms as $p[x]P$, and the system-signed arms as $p[x]S$, the primary risk-difference treatment effect will be calculated as:

$$Risk\ difference\ of\ any\ mailer\ versus\ no\ mailer: \frac{(pPP + pPS + pNP + pNS)}{4} - pC$$

$$Risk\ difference\ of\ pre\text{–}commitment\ prompt\ versus\ no\ prompt: \frac{pPP - pNP}{2} + \frac{pPS - pNS}{2}$$

$$Risk\ difference\ of\ provider\text{–}signed\ versus\ system\text{–}signed: \frac{pPP - pPS}{2} + \frac{pNP - pNS}{2}$$

Each contrast will assign equal weight across the relevant factorial cell. Relative effects will be summarized using risk ratios calculated from the standardized outcome probabilities, calculated as:

$$Risk\ ratio\ of\ any\ mailer\ versus\ no\ mailer: \frac{(pPP + pPS + pNP + pNS)/4}{pC}$$

$$Risk\ ratio\ of\ pre\text{–}commitment\ prompt\ versus\ no\ prompt: \frac{pPP + pPS}{pNP + pNS}$$

$$Risk\ ratio\ of\ provider\text{–}signed\ versus\ system\text{–}signed: \frac{pPP + pNP}{pPS + pNS}$$

The three primary comparisons address distinct research questions and will be tested separately without adjustment for multiplicity. Therefore, the familywise type I error rate across the three primary comparisons will not be controlled.

**Interaction and factor-specific effects**

To assess whether the average factorial effects conceal heterogeneity across the levels of the other factor, we will estimate the pre-commitment prompt by message-source interaction on the risk-difference scale:

$$Risk\ difference\ of\ pre\text{–}commitment\ by\ message\text{–}source\ interaction: (pPP - pNP) - (pPS - pNS)$$

We will report the interaction estimate and 95% confidence interval. To provide additional context for a potential interaction estimate, we will report the adjusted outcome probability in each of the four mailer arms and the simple effect of each factor at each level of the other factor. However, the trial is not designed to detect an interaction effect, and so interaction estimates will be interpreted based on the magnitude and the 95% confidence intervals rather than on statistical significance alone.

**Secondary Analyses**

Secondary outcomes will be analyzed using the same logistic regression model, marginal standardization, and contrasts used for the primary outcome analyses. We will also examine age and serious illness subgroups. Specifically, analyses will examine intervention effects for patients age 65 and above and below 65, and for patients included on a qualifying disease registry. For each subgroup variable, the model will include the subgroup indicator and its interactions with each of the four mailer-arm indicators.
