## Supplementary Materials: DSMB Charter for "Study protocol for a factorial randomized trial of behavioral interventions to increase health care proxy documentation among older and seriously ill adults"

### Standing DSMB Charter

Project Title:  Edward R. Roybal Centers for Translational Research in the Behavioral and Social Sciences of Aging (RFA-AG-24-006 & RFA-AG-24-007).  This project has multiple individual grants.

| Study Name | Grant Number | PI |
|---|---|---|
| Using reinforcement learning to personalize electronic health record tools to facilitate deprescribing | 2P30AG064199-06 | CHOUDHRY, NITEESH K<br><br>ZAMBRANO, JOHN |
| Behavioral Interventions Combining AI/ML and Personalized Medicine to Reduce Cardiovascular Risk | 2P30AG034532-16 | DOYLE, JOSEPH J<br><br>FADLON, ITZIK<br><br>TAI-SEALE, MING |
| Understanding Mechanisms of Action of the Tailored Activity Program for People with Dementia in Adult Day Services | 1P30AG086642-01 | GAUGLER, JOSEPH E<br><br>PARKER, LAUREN |
| Improving Screening for Primary Aldosteronism through a Comprehensive Intervention Featuring Active and Default Choice Provider Alerts | 2P30AG034546-16 | VOLPP, KEVIN G<br><br>HERMAN, DANIEL |
| A Randomized Trial to Reduce Inappropriate Prescribing to Older Adults Visiting the Emergency Department | 2P30AG024968-22 | DOCTOR, JASON N<br><br>MEEKER, DANIELLA |
| A Randomized Trial to Encourage Older Adults to Designate a Health Care Proxy | 2P30AG024968-22 | DOCTOR, JASON N<br><br>JACOBSON, MIRELLE |
| Latinos Increasing Fuerza through Exercise & Diet (LIFTED) | 2P30AG022849-21 | MARQUEZ, DAVID X<br><br>AGUINAGA, SUSAM |
| Leveraging Adult Protective Service Interaction to Offer Evidence Based Treatment for Depression in Elder Neglect/Self Neglect | 1P30AG086563-01 | PICKERING, CAROLYN E ZIMINSKI<br><br>WOOD, LEILA |
| Web-Based Support Program for AD/ADRD Caregivers of Hospitalized Veterans Living with Dementia to Prevent Elder Mistreatment during Care Transitions from Hospital to Home | 1P30AG086563-01 | PICKERING, CAROLYN E ZIMINSKI<br><br>HORSTMAN, MOLLY |
| pending | P30 AG064190 | |

| | |
|---|---|
| pending | P30 AG064198 |
| pending | P30 AG022845 |
| pending | P30 AG063786 |
| pending | P30 AG086637 |
| pending | P30 AG086635 |
| pending | P30 AG064200 |
| pending | P30 AG086562 |
| pending | P30 AG086561 |

The Data and Safety Monitoring Board (DSMB) will act in an advisory capacity to the National Institute of Aging (NIA) Director to monitor participant safety, data quality and progress of the individual grant numbers.

#### DSMB Responsibilities

The DSMB responsibilities are to:

Initial meeting

- Review the entire IRB-approved study protocol and the MOP, with regard to participant safety, recruitment, randomization, intervention, data management, quality control and analysis and the informed consent document.

- Recommend changes to the protocol and the informed consent form, when applicable.

- Identify the relevant data parameters and the format of the information to be regularly reported.

- Recommend participant recruitment be initiated after receipt of a satisfactory protocol. If the need for modifications to the protocol, the MOP, consent form, DSMP or any other study document is indicated by the DSMB and/or the NIA Program Officer (PO), the DSMB will postpone its recommendation for the initiation of participant recruitment until after the receipt of a satisfactory revised protocol(s) or other study documents.

During the study meetings

- Review masked and unmasked data. These data can be related to safety, recruitment, randomization, retention, protocol adherence, trial operations, data completeness, form completion, intervention effects, gender and minority inclusion.

- Identify needs for additional data relevant to safety issues and request these data from the study investigators.

- Propose additional analyses and periodically review developing data on safety and endpoints.

- At each meeting, consider the rationale for continuation of the study, with respect to progress of randomization, retention, protocol adherence, data management, safety issues, and outcome data (if relevant) and make a recommendation for or against the trial's continuation.

- Review and make recommendations on proposed protocol changes, and/or new protocols proposed during the trial. When the DSMBs are unblinded, the Boards may recommend to NIA to appoint a blinded working group of the DSMB to review the proposed protocol changes and make recommendations to NIA on whether to approve the requests.

- Provide advice on issues regarding data discrepancies found by the data auditing system or other sources.

- Review manuscripts of trial results if requested by the Board or the NIA PO   who may seek DSMB review of manuscripts reporting major outcomes prior to their submission for publication.

The DSMB will discharge itself from its duties when the study is complete.

#### Membership

Membership consists of persons completely independent of the investigators who have no financial, scientific or other conflict of interest with the trial. Collaborators or associates of the PI are not eligible to serve on the DSMB. Written documentation attesting to absence of conflict of interest is required and will be collected by NIA. This DSMB will consist of 12 members; split into cohort 1 and cohort 2. It is anticipated 6 of the 12 members will be invited to an individual meeting.  Members having been approved by the Director of NIA. A quorum will consist of a simple majority of voting members to include the DSMB Chair and the DSMB Statistician.

The DSMB includes experts in or representatives of the fields of:

- Gerontology, Geriatrics

- Biostatistics
- Epidemiology
- Health Policy
- Community Health
- Behavioral Mechanisms
- Digital Health
- Psychiatry/Psychology
- NIH Stage Modeling
- Psychiatry
- Ethics

Inbal Billie Nahum-Shani, PhD (cohort 1) and Belinda Borrelli, PhD (cohort 2) have been nominated by NIA to serve as the respective Chairpersons. and are responsible for facilitating the meetings, reviewing the first draft of the meeting notes with the NIA Program Official and any decision making in the case of a tie vote. The Chair and NIA Program Officer are the contact people for the DSMB. The Roybal Coordinating Center will provide the logistical management and support for the DSMB.

#### Meeting Format

Meetings of the DSMB will be held regularly (e.g., every six to nine months) at the call of NIA or the DSMB Chair. The NIA Program Officer or designee (NIA staff) will be present at every meeting. An emergency meeting of the DSMB may be called at any time by the Chair or the NIA, should participant safety questions or other unanticipated problems arise.  Refer to the individual grant Data Safety Monitoring Plan (DSMP) for meeting frequency and open and/or closed reports to be provided.

DSMB meetings will consist of open, closed and optional executive sessions, all closed to the public because discussions may address confidential participant data. The study PI and key staff members, DSMB members and NIA Program Officer and/or authorized NIA staff attend the **open sessions**. Discussions at these sessions focus on the review of the aggregate data, conduct and progress of the study, including participant accrual, protocol compliance, and problems encountered. Data by treatment group are not presented in the open session.

The **closed session** will be attended by unblinded study staff, the DSMB members and the NIA PO or designee(s). The NIA PO attends the closed and open sessions as an observer, not as a DSMB member, to answer any policy or administrative questions the DSMB members may have. The primary objective of the closed sessions is to review data by study group. To ensure participants safety and well-being, DSMBs for NIA-funded trials are required to review safety data by the actual treatment group. In many instances, safety data could also be the outcome data. Therefore, the unblinded Boards no longer review and provide recommendations to NIA on any, but safety-related protocol changes. All other protocol modifications are subject to review by the blinded working groups of the DSMBs. DSMBs’ working groups are appointed by NIA and provide their recommendations to NIA Director who makes decisions about whether to approve or decline proposed modifications.

If necessary, an **executive session** may be requested by the DSMB and will be attended only by voting DSMB members. The NIA Program Officer or designee is not permitted to attend the executive sessions.

The NIA PO, DSMB Chair or the Principal Investigator will prepare the meeting agenda that usually includes the following:

1. Welcome and introduction – study team, DSMB members, NIA and CRSS staff
2. Open session (review study protocol and its amendments, consent form, open study report, etc.) - study team, DSMB members, NIA and CRSS staff
3. Closed session (review closed session report, including unblinded data, etc.) – unblinded study statistician, DSMB members, NIA and CRSS staff
4. Executive session (optional, upon DSMB request) – DSMB members only
5. Debriefing (optional, upon DSMB request, time permitting) - study team, DSMB members, NIA and CRSS staff

The DSMB may modify its processes and procedures at any time with the approval of the NIA PO.

#### Meeting Materials

DSMB interim report templates developed by the study staff or study-specific versions of NIA report templates for both the open and closed sessions and plans for interim analyses will be reviewed and either approved at the initial DSMB meeting or changes requested. Upon DSMB request and approval by NIA, reports could be modified at any time during the study. All meeting materials should be sent by email to NIA at least 2 weeks (10 full business days, excluding holidays) prior to the meeting. NIA PO will send the reports to the DSMB.

**Part 1 - Open Session Reports:** Open session reports are individually grant based and will include administrative reports that describe participants screened, enrolled, completed, and discontinued, as well as baseline characteristics of the study population. Other general information on study status may also be presented. Listings of adverse events and serious adverse events as well as any other information requested by the DSMB may also be in the open session report, but none of the data will be presented in an unblinded manner.

**Part 2 – Closed Session Report:** Closed session reports are individually grant based and will present the same information as presented in the open session but by unblinded treatment group.

#### Reports from the DSMB

A report containing the recommendations for continuation or modification of the study will be prepared by the DSMB, NIA Project Officer or NIA contractors. The draft report will be sent to the DSMB members for review and approval not later than three weeks after the meeting. Once approved by the DSMB members, the Program Official will forward the DSMB recommendations to the Principal Investigator indicating NIA's concurrence with the report or its parts. It is the responsibility of the Principal Investigator to distribute the DSMB recommendation to all co-investigators and to ensure that copies are submitted to the IRB that reviewed and approved the study documents.

As it stated above, each meeting must include a recommendation to continue the study made by a formal DSMB majority or unanimous vote. Should the DSMB decide to issue a termination recommendation, the full vote of the DSMB is required. In the event of a split vote, majority vote will rule and a minority report should be appended. The DSMB Chair provides the tiebreaking vote in the event of a 50-50 split vote.

A recommendation to terminate the study may be made by the DSMB at any time by majority vote. If this recommendation was made during the DSMB’s Executive session, the Chair should provide notify the NIA immediately by telephone and email. After the NIA Director makes a decision about whether to accept or decline the DSMB recommendation to terminate the study, NIA PO informs the PI about the decision.

#### Miscellaneous

Refer to the individual grant DSMP for Serious Adverse Event reporting procedures to the DSMB.

#### Confidentiality

All materials, discussions and proceedings of the DSMB are completely confidential. Members and other participants in DSMB meetings are expected to maintain confidentiality.
